# Short-term drop in antibody titer after the third dose of SARS-CoV-2 BNT162b2 vaccine in adults

**DOI:** 10.1101/2022.03.07.22272028

**Authors:** Jonas Herzberg, Bastian Fischer, Heiko Becher, Ann-Kristin Becker, Human Honarpisheh, Salman Yousuf Guraya, Tim Strate, Cornelius Knabbe

**Author notes:** **Corresponding Author:** Dr. Jonas Herzberg, Krankenhaus Reinbek St. Adolf-Stift, Department of Surgery, Hamburger Str. 41, 21465 Reinbek Germany, E-Mail. Shared co-first authorship (Jonas Herzberg, Bastian Fischer).

## Abstract

Little is known about the longevity of antibodies after a third dose of BNT162b2 (BioNTech/Pfizer). Therefore, the serum antibody levels were evaluated after the third dose of BNT162b2 which dropped significantly within 11 weeks from 4155.59 ± 2373.65 BAU/ml to 2389.10 ± 1433.90 BAU/ml, *p*-value <0.001 but remained higher than after the second dose.

These data underline the positive effect of third dose of BNT162b2 but shows a rapid and significant drop of antibodies within a short span of time.

**Trial Registration:** This trial was prospectively registered in the German Clinical Trial Register (DRKS00021270).

## Introduction

The protective role of SARS-CoV-2 BNT162b2 vaccine, as introduced by BioNTech/Pfizer, has been endorsed by several studies[1,2]. The rapidly increasing number of COVID-19 cases among the previously vaccinated individuals has led to the addition of a third dose of vaccines as a booster [3,4]. The immediate immune response after the third dose has been evaluated by a wide range of clinical trials [5–8]. Till now, there is a lack of best clinical evidence about the durability and long-term impact of the immune response that is invariably induced by the third dose.

Foregoing in view, we assessed the anti-spike-IgG-antibody-titers, 4 and 11 weeks after the third dose of BNT162b2 as well as 11 weeks after the administration of the second dose of BNT162b2 within a previously elaborated study cohort.

## Methods

We included employees of the Hospital Reinbek St. Adolf-Stift, a German secondary care hospital to the ProCoV-study, established in April 2020 [9]. In the follow-up analysis, all participants with a homogeneous prime-boost-protocol, who had received three doses of BNT162b2, were invited to participate in this study. Participants with a previous self-reported SARS-CoV-2-infection within the last six months before the last blood drawn were excluded.

All participants provided a blood sample 11 weeks after the second dose of vaccinate and then 4 weeks after the third dose between November and December 2021. Subsequently, in January 2022, a follow-up blood sample was drawn 11 weeks after the third dose.

Anti-SARS-CoV-2-IgG antibodies were measured using the anti-SARS-CoV-2-assay (IgG) from Abbott (Chicago, USA) and the values were quantitatively expressed in binding-antibody-units per ml (BAU/ml) In addition, neutralizing anti-SARS-CoV-2 antibodies where detected using the NeutraLISA™ SARS-CoV-2 Neutralization Antibody Detection KIT (Euroimmun, Lübeck, Germany). The study was approved in April 2020 by the ethics committee of the medical association Schleswig-Holstein, Germany and all participants provided written and informed consent prior to inclusion. The statistical analysis was done using the IBM SPSS Statistics Version 25 (IBM Co., Armonk, NY, USA) and GraphPad Prism 9.

All variables were presented as means with standard deviation and medians with interquartile range. Categorical variables were shown as numbers with percentages. The differences between groups were analyzed using the Wilcoxon test. A *p*-value < 0.05 was considered statistically significant.

## Results

Overall, 53 participants fulfilled the inclusion criteria, of which 103 provided a blood sample 11 weeks after the third dose. Six participants were excluded from this cohost due to a reported SARS-CoV-2-infection within the last six months. The characteristics of the recruited 97 participants are shown in **Table 1**.

**Table 1:**
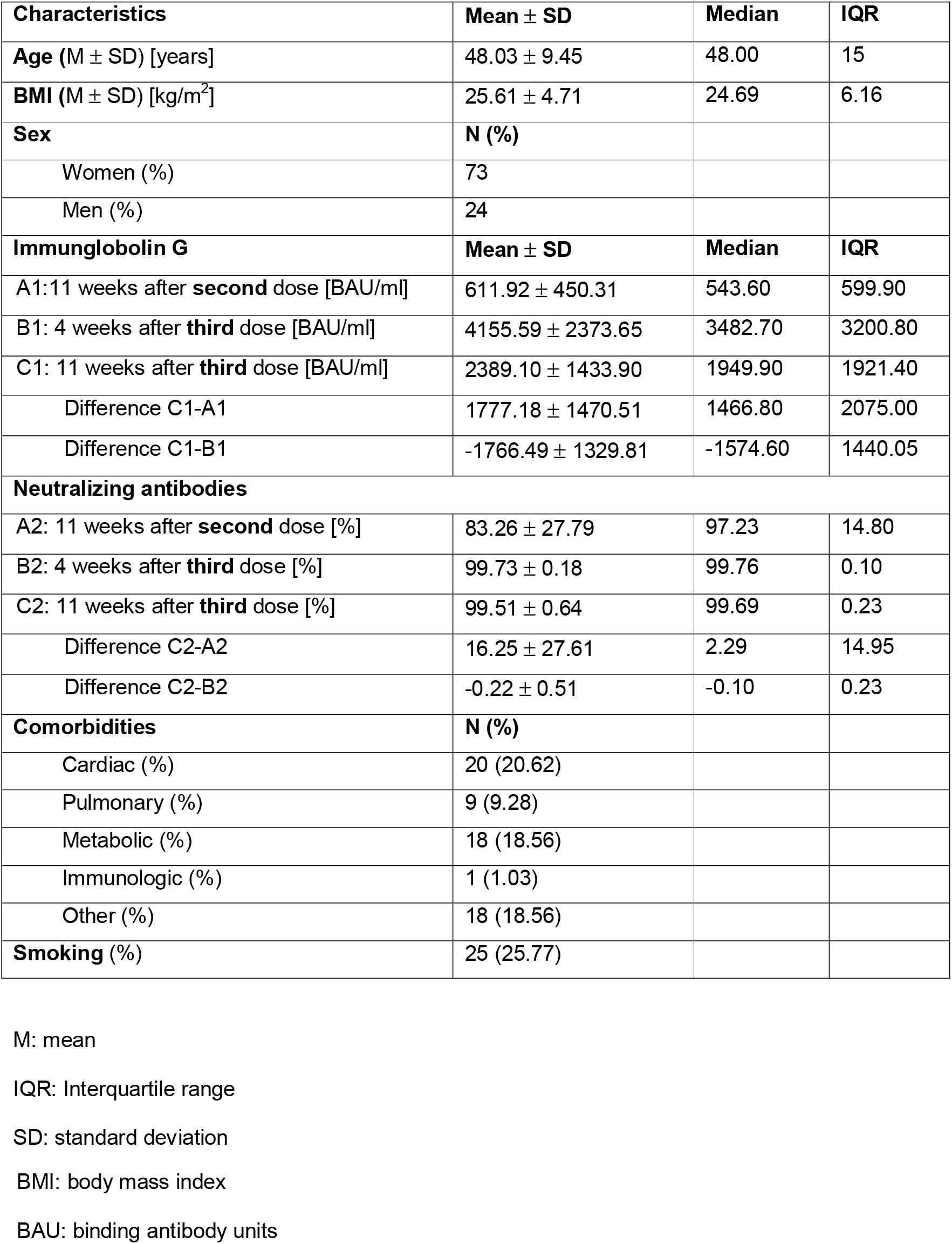
Baseline characteristics of the study cohort (N = 97)

| Characteristics | Mean ± SD | Median | IQR |
| --- | --- | --- | --- |
| **Age** (M ± SD) [years] | 48.03 ± 9.45 | 48.00 | 15 |
| **BMI** (M ± SD) [kg/m<sup>2</sup>] | 25.61 ± 4.71 | 24.69 | 6.16 |
| **Sex** | **N (%)** |  |  |
| Women (%) | 73 |  |  |
| Men (%) | 24 |  |  |
| **Immunglobolin G** | **Mean ± SD** | **Median** | **IQR** |
| A1: 11 weeks after **second** dose [BAU/ml] | 611.92 ± 450.31 | 543.60 | 599.90 |
| B1: 4 weeks after **third** dose [BAU/ml] | 4155.59 ± 2373.65 | 3482.70 | 3200.80 |
| C1: 11 weeks after **third** dose [BAU/ml] | 2389.10 ± 1433.90 | 1949.90 | 1921.40 |
| Difference C1-A1 | 1777.18 ± 1470.51 | 1466.80 | 2075.00 |
| Difference C1-B1 | -1766.49 ± 1329.81 | -1574.60 | 1440.05 |
| **Neutralizing antibodies** |  |  |  |
| A2: 11 weeks after **second** dose [%] | 83.26 ± 27.79 | 97.23 | 14.80 |
| B2: 4 weeks after **third** dose [%] | 99.73 ± 0.18 | 99.76 | 0.10 |
| C2: 11 weeks after **third** dose [%] | 99.51 ± 0.64 | 99.69 | 0.23 |
| Difference C2-A2 | 16.25 ± 27.61 | 2.29 | 14.95 |
| Difference C2-B2 | -0.22 ± 0.51 | -0.10 | 0.23 |
| **Comorbidities** | **N (%)** |  |  |
| Cardiac (%) | 20 (20.62) |  |  |
| Pulmonary (%) | 9 (9.28) |  |  |
| Metabolic (%) | 18 (18.56) |  |  |
| Immunologic (%) | 1 (1.03) |  |  |
| Other (%) | 18 (18.56) |  |  |
| **Smoking (%)** | 25 (25.77) |  |  |

M: mean
IQR: Interquartile range
SD: standard deviation
BMI: body mass index
BAU: binding antibody units

Later, 11 weeks after the second dose, 93 participants (95.9%) showed a positive antibody response with a mean titer of 611.92 ± 450.31 BAU/ml. our Four weeks after the third dose, all participants showed a seropositive result and remained seropositive 11 weeks after the third dose.

A comparison of the antibody titer 11 weeks after second dose with those 11 weeks after the third dose showed a significant rise in antibody titer that was induced by the third dose (611.92 ± 450.31 vs. 2389.10 ± 1433.90, *p-value* < 0.001). A corresponding rise in the neutralizing antibodies (83.26 ± 27.79 vs. 99.51 ± 0.64, *p-value* < 0.001) was also reported.

Interestingly, the mean IgG-titer four weeks after the third dose was found to be 4155.59 ± 2373.65 BAU/ml which dropped significantly to 2389.10 ± 1433.90 BAU/ml 11 weeks after the third dose (*p-value* < 0.001) (**Figure 1A**). This drop was also noted in the neutralizing antibodies titer, which also showed a significant decrease in the weeks after the third dose but remained at a high level (99.73% ± 0.18% vs. 99.51, *p-value* < 0.001) (**Figure 1B**).

**Figure 1:**
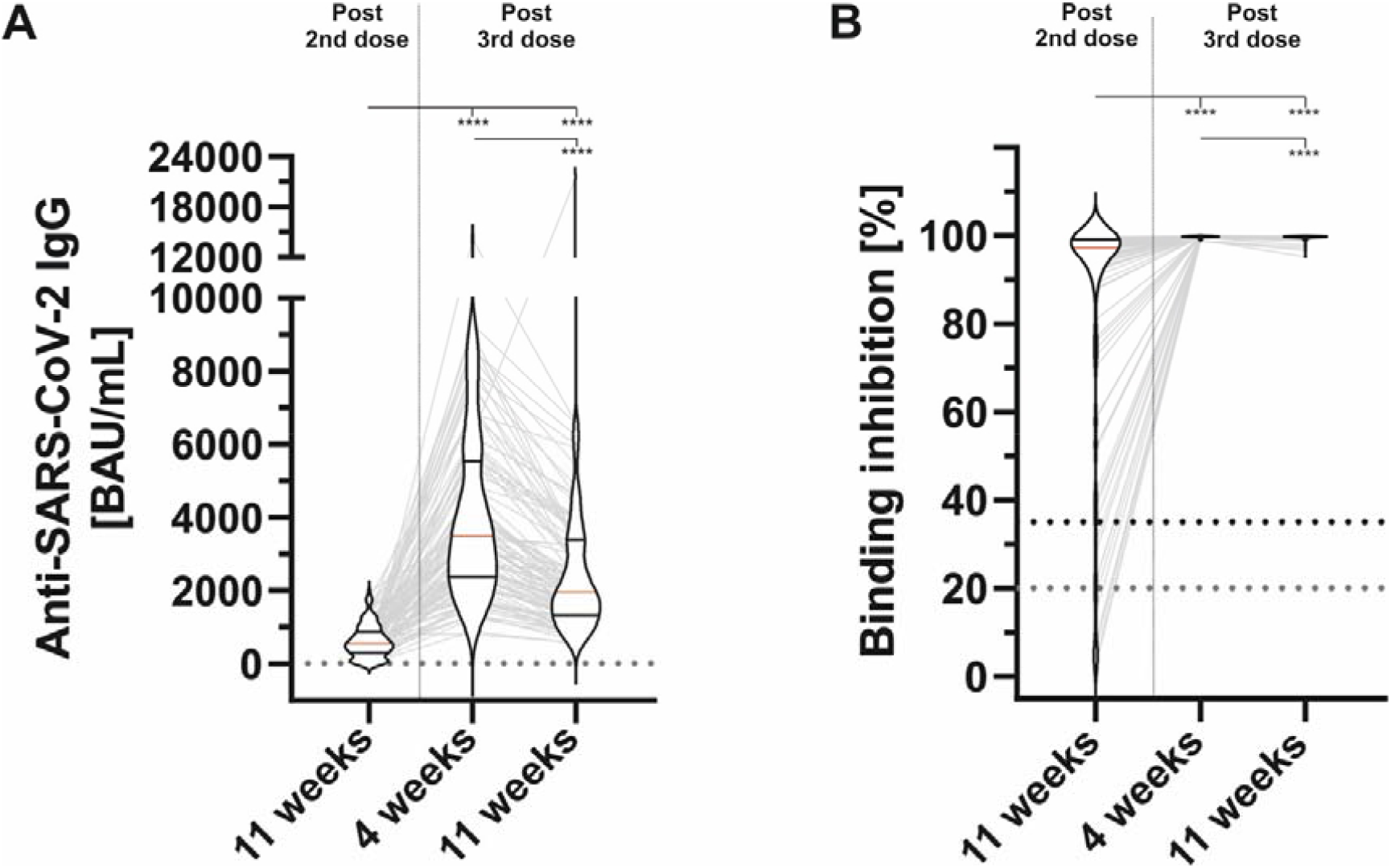
The vaccine induced serological immune response at different timepoints post second- and third doses of BNT162b2. A: Anti-SARS-CoV-2-IgG at different timepoints (left: 11 weeks post second vaccine dose, right: 4- and 11 weeks after third vaccine dose, respectively). B: Binding inhibition at different timepoints (left: 11 weeks post second vaccine dose, right: 4- and 11 weeks after third vaccine dose, respectively). Horizontal lines within plotted data show the median (red line) and interquartile ranges. ****p < 0.0001 (Mann-Whitney U-test).

## Discussion

Our study demonstrates a significant rise in the binding-antibody titer and the neutralizing antibodies following the third dose as compares to the serological response to the second dose of BNT162b2. On the other hand, our data has reported a rapid and significant drop in the binding-antibody titers and neutralizing antibodies in a short term (within 7 weeks) after third vaccination, though the titers remain high.

Till now, little is known about the sustained and long-term impact of the vaccination-induced immune response. Despite a small cohort size of our study, several other studies have shown positive inductive immunological effect of a third dose of BNT162b2 [5,6,8]. Our study provides an original clinical data about the antibody persistence of the second- and third dose during a short-term follow-up. While persistence of the humoral immunity is much stronger after the administration of the third dose of BNT162b2, the dropping antibody titers following the third dose signals the potential indication of a fourth dose of vaccine. This possibility has been recently suggested by some recent reports [10,11].

## Data Availability

All data produced in the present study are available upon reasonable request to the authors.

## Limitations

This study is limited by a small sample size and by the lack of cellular immunity testing. Further studies are needed to evaluate the infection rate depending on several antibody titers which can detect potential measurable thresholds.

## Conclusion

This cohort study provides first human-based clinical data about the longevity of the humoral immune response induced by a third dose of BNT162b2 against COVID-19. This data may potentially facilitate to determine the indication of an additional fourth dose of anti-COVID-19-vaccine.

## Notes

### Competing Interest Statement

The authors have declared no competing interest.

### Funding Statement

This study did not receive any funding.

### Author Declarations

Ethics committee of the medical association Schleswig-Holstein gave ethical approval for this work.

## References

1. Sahin U, Muik A, Derhovanessian E, Vogler I, Kranz LM, Vormehr M, et al. COVID-19 vaccine BNT162b1 elicits human antibody and TH1 T cell responses. Nature. 2020 Oct 22;586(7830):594–9. http://www.nature.com/articles/s41586-020-2814-7

2. Polack FP, Thomas SJ, Kitchin N, Absalon J, Gurtman A, Lockhart S, et al. Safety and Efficacy of the BNT162b2 mRNA Covid-19 Vaccine. N Engl J Med. 2020;383(27):2603–15. http://www.ncbi.nlm.nih.gov/pubmed/33301246

3. Bar-On YM, Goldberg Y, Mandel M, Bodenheimer O, Freedman L, Kalkstein N, et al. Protection of BNT162b2 Vaccine Booster against Covid-19 in Israel. N Engl J Med. 2021 Oct 7;385(15):1393–400. http://www.nejm.org/doi/10.1056/NEJMoa2114255

4. Saciuk Y, Kertes J, Shamir Stein N, Ekka Zohar A. Effectiveness of a third dose of BNT162b2 mRNA vaccine. J Infect Dis. 2021 Nov 2; http://www.ncbi.nlm.nih.gov/pubmed/34726239

5. Eliakim-Raz N, Leibovici-Weisman Y, Stemmer A, Ness A, Awwad M, Ghantous N, et al. Antibody Titers Before and After a Third Dose of the SARS-CoV-2 BNT162b2 Vaccine in Adults Aged ≥60 Years. JAMA. 2021 Dec 7;326(21):2203. https://jamanetwork.com/journals/jama/fullarticle/2786096

6. Benotmane I, Gautier G, Perrin P, Olagne J, Cognard N, Fafi-Kremer S, et al. Antibody Response After a Third Dose of the mRNA-1273 SARS-CoV-2 Vaccine in Kidney Transplant Recipients With Minimal Serologic Response to 2 Doses. JAMA. 2021 Sep 21;326(11):1063. https://jamanetwork.com/journals/jama/fullarticle/2782538

7. Ducloux D, Colladant M, Chabannes M, Yannaraki M, Courivaud C. Humoral response after 3 doses of the BNT162b2 mRNA COVID-19 vaccine in patients on hemodialysis. Kidney Int. 2021 Sep 1;100(3):702–4. http://www.ncbi.nlm.nih.gov/pubmed/34216675

8. Stumpf J, Tonnus W, Paliege A, Rettig R, Steglich A, Gembardt F, et al. Cellular and Humoral Immune Responses After 3 Doses of BNT162b2 mRNA SARS-CoV-2 Vaccine in Kidney Transplant. Transplantation. 2021 Nov 22;105(11):e267–9. http://www.transplantjournal.com

9. Herzberg J, Vollmer T, Fischer B, Becher H, Becker A-K, Sahly H, et al. A Prospective Sero-epidemiological Evaluation of SARS-CoV-2 among Health Care Workers in a German Secondary Care Hospital. Int J Infect Dis. 2020 Oct 16;0(0). http://www.ncbi.nlm.nih.gov/pubmed/33075538

10. Standing Committee on Vaccination. Press release STIKO [Internet]. https://www.rki.de/DE/Content/Kommissionen/STIKO/Empfehlungen/PM_2022-02-03.html

11. Burki TK. Fourth dose of COVID-19 vaccines in Israel. Lancet Respir Med. 2022 Jan 11;10(2):e19. http://www.ncbi.nlm.nih.gov/pubmed/35030317

